# Cumulative incidence of SARS-CoV-2 infections among adults in Georgia, USA, August-December 2020

**DOI:** 10.1101/2021.05.06.21256407

**Authors:** Allison T. Chamberlain, Kathleen E. Toomey, Heather Bradley, Eric W. Hall, Mansour Fahimi, Benjamin A. Lopman, Nicole Luisi, Travis Sanchez, Cherie Drenzek, Kayoko Shioda, Aaron J Siegler, Patrick Sean Sullivan

## Abstract

**Background:** Reported COVID-19 cases underestimate the true number of SARS-CoV-2 infections. Data on all infections, including asymptomatic infection, are needed to guide state testing and prevention programs. To minimize biases in estimates from seroprevalence surveys and reported cases, we conducted a state-wide probability survey of Georgia households and estimated cumulative incidence of SARS-CoV-2 infections adjusted for antibody waning.

**Methods:** From August to December 2020, we mailed kits to self-collect specimens (nasal swabs and blood spots) to a random sample of Georgia addresses. One randomly-selected adult household member completed a survey and returned specimens for virus and antibody testing. We estimated cumulative incidence of SARS-CoV-2 infections adjusted for waning antibodies, reported fraction, and infection fatality ratio (IFR). Differences in seropositivity among demographic, geographic and clinical subgroups were explored with weighted prevalence ratios (PR).

**Results:** Among 1,370 Georgia adult participants, adjusted cumulative incidence of SARS-CoV-2 was 16.1% (95% credible interval (CrI): 13.5-19.2%) as of November 16, 2020. The reported fraction was 26.6% and IFR was 0.78%. Non-Hispanic Black (PR: 2.03, CI 1.0, 4.1) and Hispanic adults (PR: 1.98, CI 0.74, 5.31) were more likely than non-Hispanic White adults to be seropositive. Seropositivity in metropolitan Atlanta’s Fulton and DeKalb counties was similar to seropositivity elsewhere in Georgia (7.8% vs. 8.8%).

**Conclusions:** As of mid-November 2020, one in 6 adults in Georgia had been infected with SARS-CoV-2. The scope of the COVID-19 epidemic in Georgia is likely substantially underestimated by reported cases.

**Main point:** Using data from a probability survey of households in Georgia, USA, we estimated that 1.3 million adults aged ≥18 years experienced SARS-CoV-2 infections by November 16, 2020, of whom 1 in 4 were reported and of whom 0.78% died.

## Introduction

Like many states In the United States, Georgia has experienced substantial morbidity and mortality due to COVID-19. Comprehensive, unbiased estimates of the extent of SARS-CoV-2 infections in Georgia are challenging because not all people who are infected have symptoms, and not all people who are symptomatic get tested. Although Georgia’s robust testing efforts have diagnosed over one million individuals(*1*), no scientifically rigorous estimate of how many Georgias have been infected with SARS-CoV-2 exists. Seroprevalence studies conducted from remnant samples in clinical settings (e.g., dialysis centers and other settings in which specimens are collected for routine screening or clinical management) can detect people who have been infected, but such studies can have biased data if they are not representative of the general population and because antibodies can become undetectable over time (“antibody waning”).(*2*)

For Georgia, ascertaining the total number of people who have been infected has implications for understanding the impact of COVID-19 to date and for reaching herd immunity. Having these data also can support and inform vaccination strategies. We describe findings from the COVIDVu Georgia study, a state-specific seroprevalence survey conducted among a probability-based sample of Georgia households from August to December 2020 to develop a representative estimate of the cumulative incidence of SARS-CoV-2 infection among Georgia’s adult population after adjusting for antibody waning.

## Methods

### Sampling

Our sampling methods have been previously described as part of the national COVIDVu study.(*3*) We used a national address-based household sample derived from the USPS Computerized Delivery Sequence File, which contains about 130 million residential addresses and covers all residential delivery points in the US. This sampling frame has been used in numerous health research studies.(*4*–*6*) To achieve a total sample of 1,400 responding households from Georgia, 12,894 addresses were shipped COVIDVu study materials (Figure 1). Analogous to our national study, we oversampled households in census tracts with >50% Black residents and households with surnames likely to represent Hispanic ethnicity to overcome differentially low early response rates by Black and Hispanic persons.(*4*) We oversampled Fulton and Dekalb counties to facilitate estimation of seroprevalence in the City of Atlanta.

**Figure 1.**
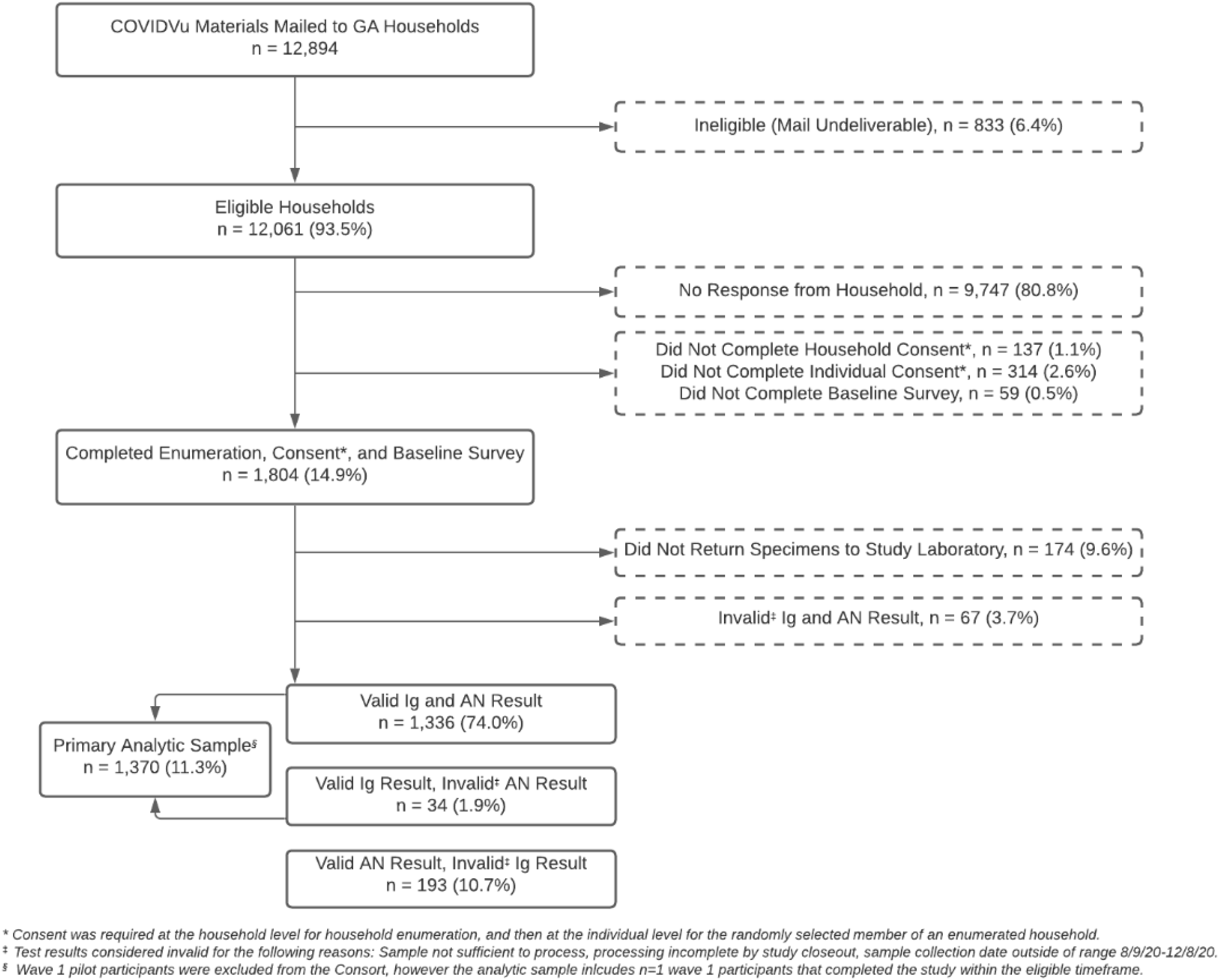
Consort diagram for a national household probability sample of US households to estimate the cumulative incidence of SARS-CoV-2 infection in Georgia, 2020.

### Survey and Laboratory Procedures

One adult ≥18 years in each household listed household members by gender and age, and an adult household member was then randomly selected for participation by the electronic data system. Following an online consent procedure, participants completed a behavioral survey with domains including demographics, comorbidities, and symptoms; the survey instrument has been previously published.(*3*) Participants self-collected an anterior nares (AN) swab and a dried blood spot (DBS) card, a method we previously validated based on clinician observation of specimen collection and laboratorian assessment of specimen quality.(*7, 8*) Specimens were returned to a central laboratory with a prepaid mailer.(*8*) PCR testing of AN swabs used the Thermo EUA Version 2 kit (Thermo Fisher Scientific, Waltham, MA). Antibody testing of DBS specimens used the BioRad Platelia Total Antibody test that targets the nucleocapsid protein (i.e., IgA, IgM, IgG; BioRad, Hercules, California). Testing protocols were validated under CLIA/CAP protocols for the development of Laboratory Developed Tests. Further detail has been previously described, including approaches taken to quantify the direction and magnitude of potential biases associated with antibody waning.(*3*) Participants in the oversample were provided with a $100 electronic gift card incentive and all other participants were provided with a $40 electronic gift card incentive. The COVIDVu study was approved by the Emory University Institutional Review Board (STUDY00000695).

### Sample Weights

We developed three sets of sample weights to allow for estimation of key parameters representing non-institutionalized and housed adults in three areas: Georgia, Fulton/Dekalb counties, and all other counties in Georgia. Each set of weights was developed using the same method as previously described. In brief, hierarchical hot deck imputation(*9*) was performed to ensure no participants were missing data for key variables needed for weighting such as gender, education, race, ethnicity, and marital status that each had less than 3% missingness. Design weights, adjusted with Classification And Regression Tree (CART) analysis for differential non-response, were developed to facilitate population inference. A raking procedure aligned weighted distributions to the observed distributions from the Census along the lines including age, race-ethnicity, education, and income.(*10*) To address outlier weights, those at the 99th percentile of each side of the distribution were trimmed. Additional detail on the weighting process can be found in our protocol paper.*(3)*

We estimated weighted seroprevalence and 95% confidence limits of total Ig for the entire sample and by demographic factors and reported pre-existing comorbidities, month of sampling, and symptoms. To identify significant differences in seroprevalence among groups, we estimated prevalence ratios (PRs) and corresponding 95% Wilson Modified confidence intervals (CIs) using weighted logistic regression. All analyses were conducted in SAS v9.4 and SUDAAN.

### Georgia SARS-CoV-2 cumulative incidence, IFR, and reported fraction

Given the considerable evidence from population-based surveys that SARS-CoV-2 antibodies wane over time to levels below detection by numerous laboratory tests (*11*–*13*), our analysis includes a Bayesian model that accounts for waning.(*14*) By accounting for (1) the time between infection and seroconversion to detectable antibodies, (2) the time between seroconversion and seroreversion to undetectable antibody test results, and (3) the time from symptom onset to death, the model estimates IFR and cumulative incidence of SARS-CoV-2 based on the Georgia weighted seroprevalence estimate from this study as well as the reported daily counts of COVID-19 associated deaths. The model applies cumulative density functions for the time from seroconversion to seroreversion, estimated by a previous study*(14)* to adjust for antibody waning. Cumulative incidence is calculated from the total number of modeled infections since the beginning of the epidemic until the median specimen collection date of our sample (November 16, 2020). This cumulative incidence also serves as the denominator for the infection fatality ratio (IFR). The ratio of the cases reported to cumulative incidence cases (the reported fraction) was developed from confirmed PCR+ cases in Georgia as of November 16, 2020 using data for adults ≥18 years from the Georgia Department of Public Health’s public use dataset.(*15*)

## Results

### Study sample

A total of 12,894 household addresses in Georgia were selected and mailed study materials from July-October 2020 (Figure 1). Of these, 6.4% (n=833) were unable to receive mail and excluded from the sample. Behavioral surveys were completed by 14.9% (n=1,804) households. A total of 11.3% (n=1,370) of sampled households completed a behavioral survey and returned a valid specimen for antibody testing during the study period of August 9-December 8, 2020 (Table 1). Of participating households, 43% (n=585) were in the oversampled area of Fulton/Dekalb and 57% (n=785) were from other counties in Georgia.

**Table 1.** SARS-CoV-2 serology and total immunoglobulin (IgA, IgM, or IgG) viral detection results for a probability sample of 1,370 households and weighted results, compared to the population aged ≥18 years, Georgia, 2020.

| Characteristic | Sample |  | Weighted Sample |  | GA Population<sup>1</sup> |  |
| --- | --- | --- | --- | --- | --- | --- |
|  | N | Column % | Weighted N | Column % | n | Column % |
| Overall | 1370 | 100 | 7,981,539 | 100 | 8,113,542 | 100 |
| Sex |  |  |  |  |  |  |
| Male | 523 | 38.2 | 3,754,083 | 47.0 | 3,886,408 | 47.9 |
| Female | 847 | 61.8 | 4,227,457 | 53.0 | 4,227,134 | 52.1 |
| Race/ethnicity |  |  |  |  |  |  |
| Hispanic | 91 | 6.6 | 596,425 | 7.5 | 673,103 | 8.3 |
| Non-Hispanic White | 747 | 54.5 | 4,395,985 | 55.1 | 4,485,895 | 55.3 |
| Non-Hispanic Black | 483 | 35.3 | 2,502,158 | 31.3 | 2,558,139 | 31.5 |
| Non-Hispanic Asian | 33 | 2.4 | 333,509 | 4.2 | 374,149 | 4.6 |
| Non-Hispanic Other | 16 | 1.2 | 153,463 | 1.9 | 22,256 | 0.3 |
| Age |  |  |  |  |  |  |
| 18-34 years | 313 | 22.8 | 2,394,183 | 30.0 | 2,508,449 | 30.9 |
| 35-44 years | 235 | 17.2 | 1,393,889 | 17.5 | 1,380,954 | 17.0 |
| 45-54 years | 229 | 16.7 | 1,393,699 | 17.5 | 1,399,652 | 17.3 |
| 55-64 years | 272 | 19.9 | 1,279,288 | 16.0 | 1,307,533 | 16.1 |
| 65+ years | 321 | 23.4 | 1,520,481 | 19.0 | 1,516,954 | 18.7 |

\*2019 Bridged-Race Estimates (NVSS).
N, total participants. Weighted N: sum of the weights of participants.

### Serology and PCR results unadjusted for antibody waning

The weighted seroprevalence in Georgia was 8.6% (95% CI: 6.3 - 11.8%), representing the period prevalence of detectable antibodies for August 9 - December 8, 2020 (Table 2). This suggests that 687,450 out of 8,113,542 adults in Georgia had prevalent anti-SARS-CoV-2 Ig at the time they provided a sample. Unweighted, a total of 7.2% of all specimens tested (99/1,370) were reactive for total Ig.

**Table 2.**
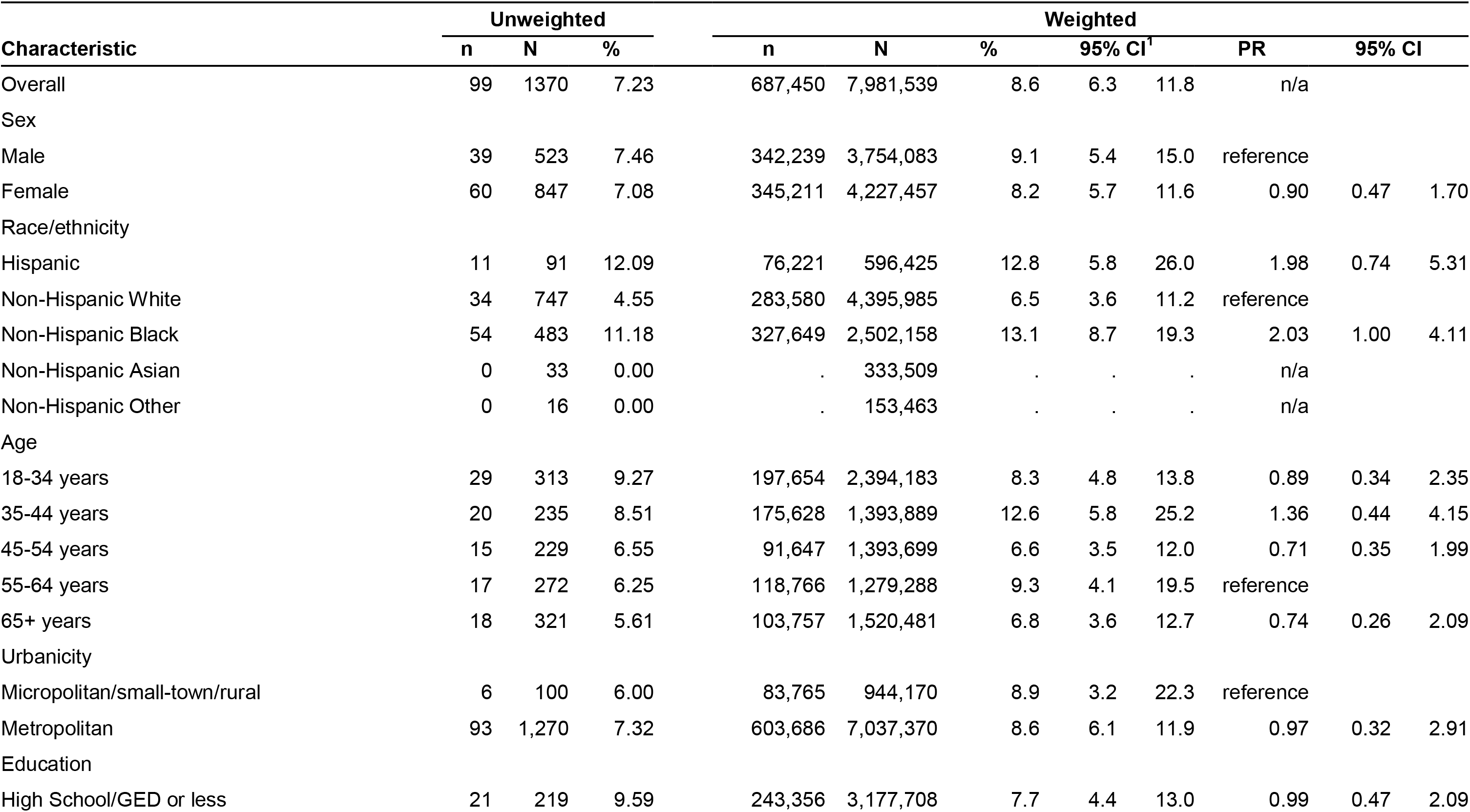
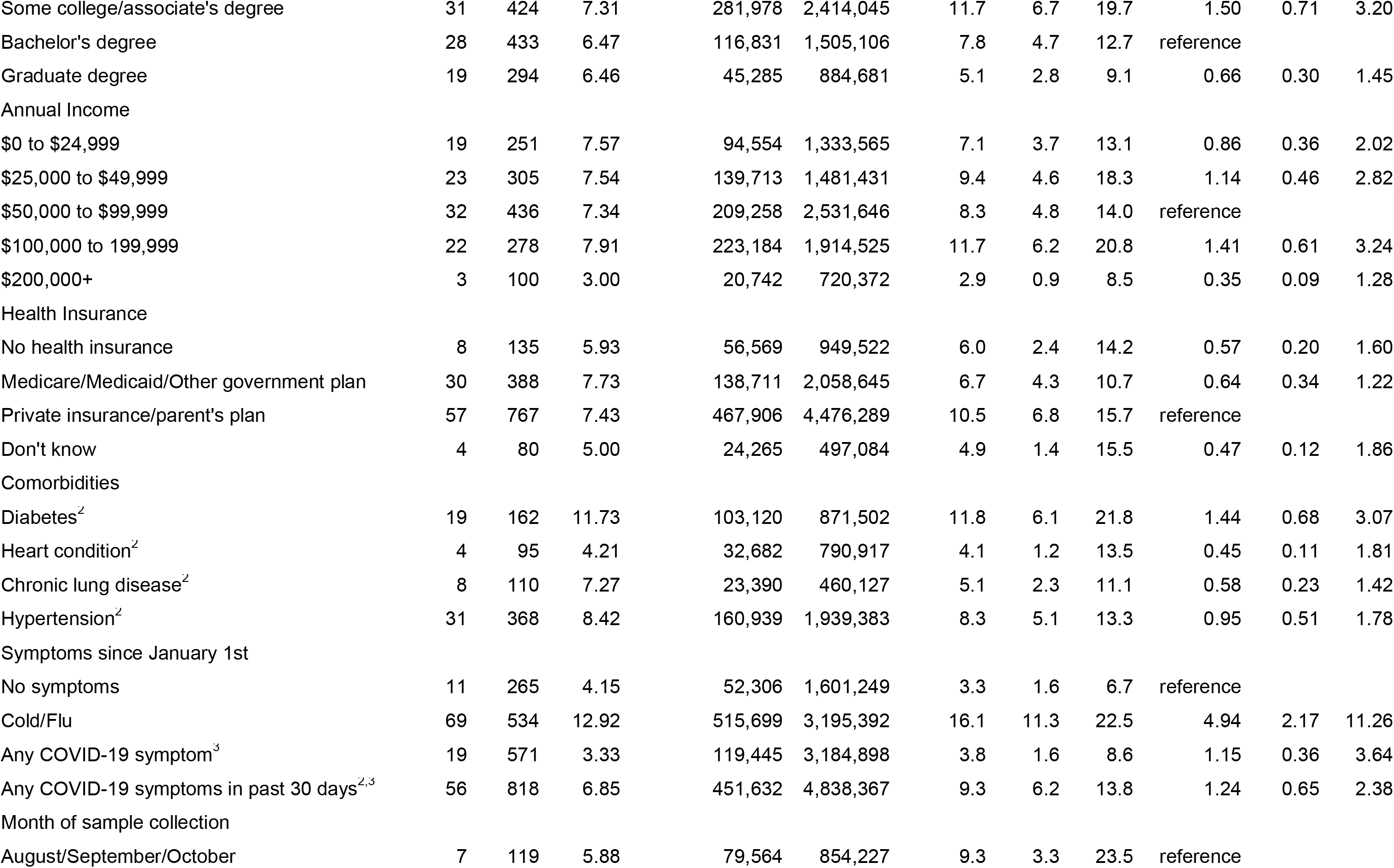
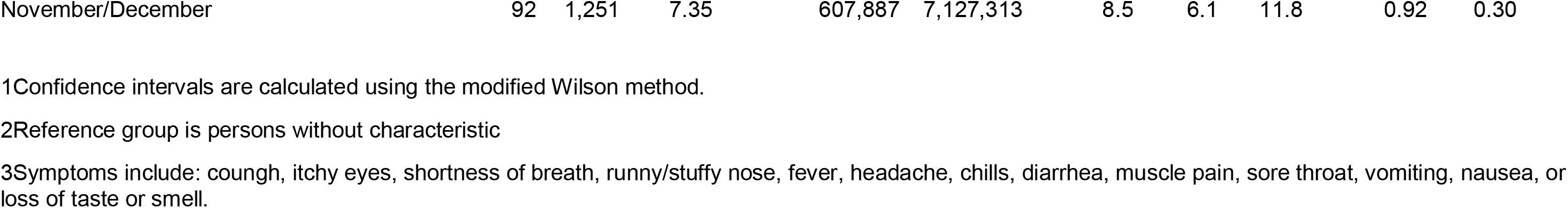
Unweighted and weighted SARS-CoV-2 antibody prevalence for a probability sample of 1,370 households and weighted results and prevalence ratios for persons aged ≥18 years, Georgia, 2020.

| Characteristic | Unweighted |  |  | Weighted |  |  |  |  |  |  |  |
| --- | --- | --- | --- | --- | --- | --- | --- | --- | --- | --- | --- |
|  | n | N | % | n | N | % | 95% CI<sup>1</sup> |  | PR | 95% CI |  |
| Overall | 99 | 1370 | 7.23 | 687,450 | 7,981,539 | 8.6 | 6.3 | 11.8 | n/a |  |  |
| Sex |  |  |  |  |  |  |  |  |  |  |  |
| Male | 39 | 523 | 7.46 | 342,239 | 3,754,083 | 9.1 | 5.4 | 15.0 | reference |  |  |
| Female | 60 | 847 | 7.08 | 345,211 | 4,227,457 | 8.2 | 5.7 | 11.6 | 0.90 | 0.47 | 1.70 |
| Race/ethnicity |  |  |  |  |  |  |  |  |  |  |  |
| Hispanic | 11 | 91 | 12.09 | 76,221 | 596,425 | 12.8 | 5.8 | 26.0 | 1.98 | 0.74 | 5.31 |
| Non-Hispanic White | 34 | 747 | 4.55 | 283,580 | 4,395,985 | 6.5 | 3.6 | 11.2 | reference |  |  |
| Non-Hispanic Black | 54 | 483 | 11.18 | 327,649 | 2,502,158 | 13.1 | 8.7 | 19.3 | 2.03 | 1.00 | 4.11 |
| Non-Hispanic Asian | 0 | 33 | 0.00 | . | 333,509 | . | . | . | n/a |  |  |
| Non-Hispanic Other | 0 | 16 | 0.00 | . | 153,463 | . | . | . | n/a |  |  |
| Age |  |  |  |  |  |  |  |  |  |  |  |
| 18-34 years | 29 | 313 | 9.27 | 197,654 | 2,394,183 | 8.3 | 4.8 | 13.8 | 0.89 | 0.34 | 2.35 |
| 35-44 years | 20 | 235 | 8.51 | 175,628 | 1,393,889 | 12.6 | 5.8 | 25.2 | 1.36 | 0.44 | 4.15 |
| 45-54 years | 15 | 229 | 6.55 | 91,647 | 1,393,699 | 6.6 | 3.5 | 12.0 | 0.71 | 0.35 | 1.99 |
| 55-64 years | 17 | 272 | 6.25 | 118,766 | 1,279,288 | 9.3 | 4.1 | 19.5 | reference |  |  |
| 65+ years | 18 | 321 | 5.61 | 103,757 | 1,520,481 | 6.8 | 3.6 | 12.7 | 0.74 | 0.26 | 2.09 |
| Urbanicity |  |  |  |  |  |  |  |  |  |  |  |
| Micropolitan/small-town/rural | 6 | 100 | 6.00 | 83,765 | 944,170 | 8.9 | 3.2 | 22.3 | reference |  |  |
| Metropolitan | 93 | 1,270 | 7.32 | 603,686 | 7,037,370 | 8.6 | 6.1 | 11.9 | 0.97 | 0.32 | 2.91 |
| Education |  |  |  |  |  |  |  |  |  |  |  |
| High School/GED or less | 21 | 219 | 9.59 | 243,356 | 3,177,708 | 7.7 | 4.4 | 13.0 | 0.99 | 0.47 | 2.09 |
| Some college/associate's degree | 31 | 424 | 7.31 | 281,978 | 2,414,045 | 11.7 | 6.7 | 19.7 | 1.50 | 0.71 | 3.20 |
| Bachelor's degree | 28 | 433 | 6.47 | 116,831 | 1,505,106 | 7.8 | 4.7 | 12.7 | reference |  |  |
| Graduate degree | 19 | 294 | 6.46 | 45,285 | 884,681 | 5.1 | 2.8 | 9.1 | 0.66 | 0.30 | 1.45 |
| Annual Income |  |  |  |  |  |  |  |  |  |  |  |
| \$0 to \$24,999 | 19 | 251 | 7.57 | 94,554 | 1,333,565 | 7.1 | 3.7 | 13.1 | 0.86 | 0.36 | 2.02 |
| \$25,000 to \$49,999 | 23 | 305 | 7.54 | 139,713 | 1,481,431 | 9.4 | 4.6 | 18.3 | 1.14 | 0.46 | 2.82 |
| \$50,000 to \$99,999 | 32 | 436 | 7.34 | 209,258 | 2,531,646 | 8.3 | 4.8 | 14.0 | reference | | |
| \$100,000 to 199,999 | 22 | 278 | 7.91 | 223,184 | 1,914,525 | 11.7 | 6.2 | 20.8 | 1.41 | 0.61 | 3.24 |
| \$200,000+ | 3 | 100 | 3.00 | 20,742 | 720,372 | 2.9 | 0.9 | 8.5 | 0.35 | 0.09 | 1.28 |
| Health Insurance |  |  |  |  |  |  |  |  |  |  |  |
| No health insurance | 8 | 135 | 5.93 | 56,569 | 949,522 | 6.0 | 2.4 | 14.2 | 0.57 | 0.20 | 1.60 |
| Medicare/Medicaid/Other government plan | 30 | 388 | 7.73 | 138,711 | 2,058,645 | 6.7 | 4.3 | 10.7 | 0.64 | 0.34 | 1.22 |
| Private insurance/parent's plan | 57 | 767 | 7.43 | 467,906 | 4,476,289 | 10.5 | 6.8 | 15.7 | reference |  |  |
| Don't know | 4 | 80 | 5.00 | 24,265 | 497,084 | 4.9 | 1.4 | 15.5 | 0.47 | 0.12 | 1.86 |
| Comorbidities |  |  |  |  |  |  |  |  |  |  |  |
| Diabetes<sup>2</sup> | 19 | 162 | 11.73 | 103,120 | 871,502 | 11.8 | 6.1 | 21.8 | 1.44 | 0.68 | 3.07 |
| Heart condition<sup>2</sup> | 4 | 95 | 4.21 | 32,682 | 790,917 | 4.1 | 1.2 | 13.5 | 0.45 | 0.11 | 1.81 |
| Chronic lung disease<sup>2</sup> | 8 | 110 | 7.27 | 23,390 | 460,127 | 5.1 | 2.3 | 11.1 | 0.58 | 0.23 | 1.42 |
| Hypertension<sup>2</sup> | 31 | 368 | 8.42 | 160,939 | 1,939,383 | 8.3 | 5.1 | 13.3 | 0.95 | 0.51 | 1.78 |
| Symptoms since January 1st |  |  |  |  |  |  |  |  |  |  |  |
| No symptoms | 11 | 265 | 4.15 | 52,306 | 1,601,249 | 3.3 | 1.6 | 6.7 | reference |  |  |
| Cold/Flu | 69 | 534 | 12.92 | 515,699 | 3,195,392 | 16.1 | 11.3 | 22.5 | 4.94 | 2.17 | 11.26 |
| Any COVID-19 symptom<sup>3</sup> | 19 | 571 | 3.33 | 119,445 | 3,184,898 | 3.8 | 1.6 | 8.6 | 1.15 | 0.36 | 3.64 |
| Any COVID-19 symptoms in past 30 days<sup>2,3</sup> | 56 | 818 | 6.85 | 451,632 | 4,838,367 | 9.3 | 6.2 | 13.8 | 1.24 | 0.65 | 2.38 |
| Month of sample collection |  |  |  |  |  |  |  |  |  |  |  |
| August/September/October | 7 | 119 | 5.88 | 79,564 | 854,227 | 9.3 | 3.3 | 23.5 | reference |  |  |

|  |  |  |  |  |  |  |  |  |  |  |
| --- | --- | --- | --- | --- | --- | --- | --- | --- | --- | --- |
| November/December | 92 | 1,251 | 7.35 | 607,887 | 7,127,313 | 8.5 | 6.1 | 11.8 | 0.92 | 0.30 |

1Confidence intervals are calculated using the modified Wilson method.
2Reference group is persons without characteristic
3Symptoms include: coughh, itchy eyes, shortness of breath, runny/stuffy nose, fever, headache, chills, diarrhea, muscle pain, sore throat, vomiting, nausea, or loss of taste or smell.

### Associations with prevalence of antibody response

For the state of Georgia, the weighted seroprevalence was two times higher for Black, non-Hispanic participants than for White, non-Hispanic participants (Table 2). A non-significant effect of similar magnitude was observed for Hispanic participants relative to White, non- Hispanic participants. Those reporting cold or flu-like symptoms after January 1, 2020 were nearly 5 times more likely than those without systems to be seropositive. Among those who were seropositive, 66/99 (weighted percent: 75%, 95% CI: 58%-86%) reported cold or flu symptoms since January 1, 2020. There were no observed differences in seroprevalence by education, income, or urbanicity.

For Fulton and DeKalb counties, point estimates of disparities in seroprevalence by race were higher than in the state as a whole, but not statistically significant (Table 3). Antibody prevalence for residents in Fulton and DeKalb counties (7.8%; CI: 5.1, 11.7) was similar to prevalence in other parts of Georgia (8.8%; CI: 6.1, 12.6). Experiencing cold or flu-like systems since the beginning of 2020 was the only variable significantly associated with seropositivity among participants not residing in Fulton or DeKalb (PR=5.2; CI: 2.0, 13.8) (Table 4).

**Table 3.**
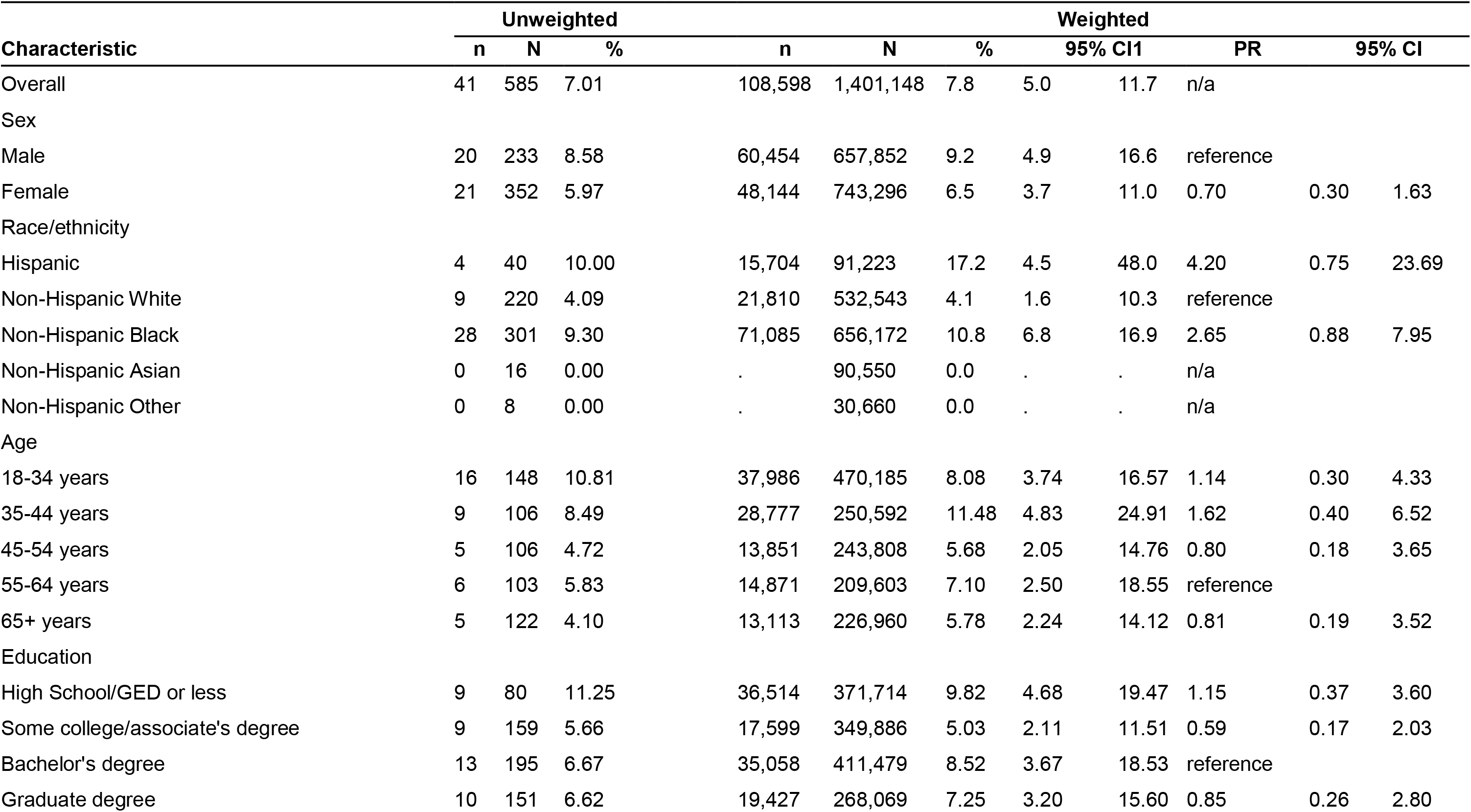
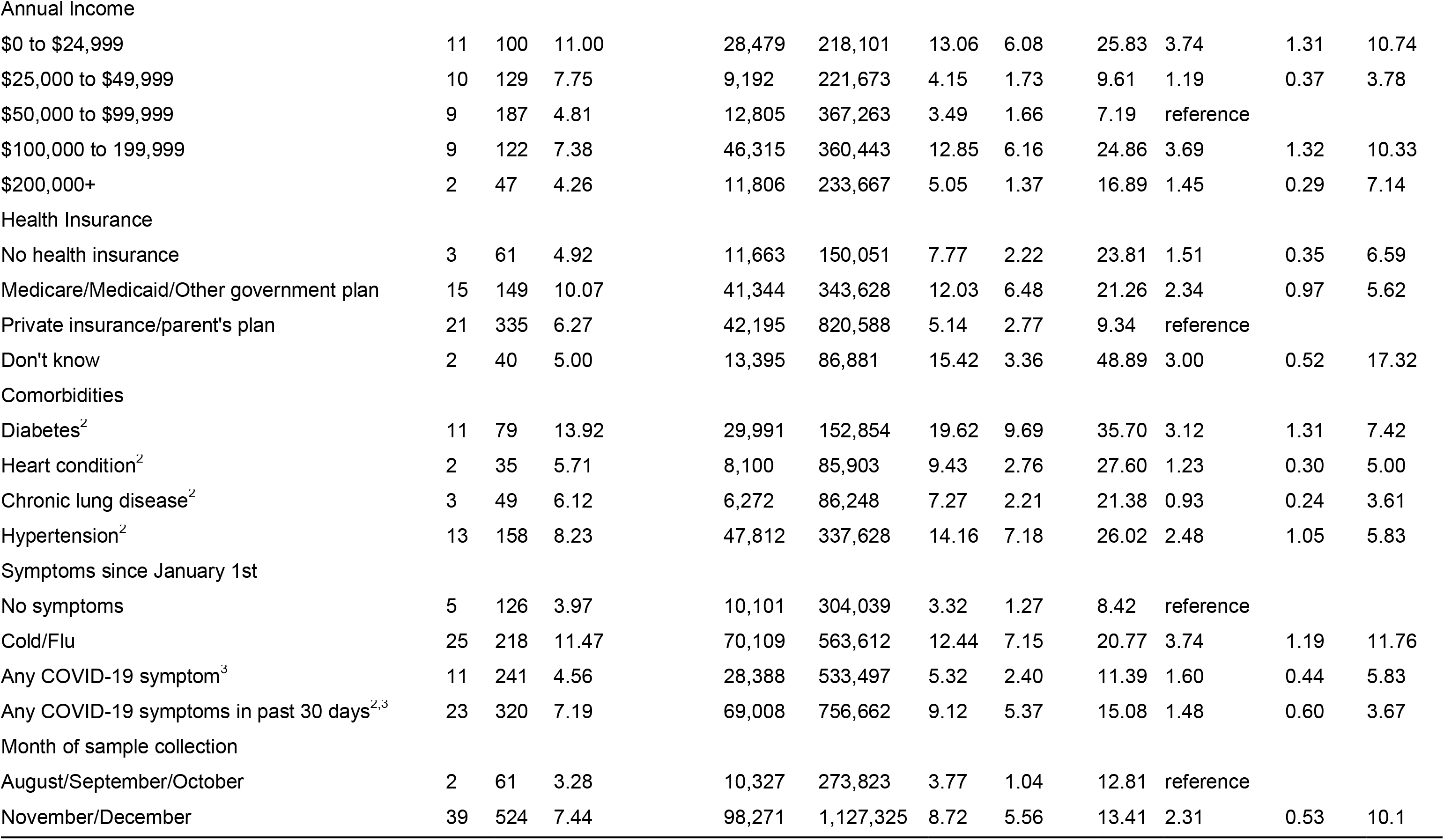
Unweighted and weighted SARS-CoV-2 antibody prevalence for a probability sample of 585 households and weighted results and prevalence ratios for persons aged ≥18 years, Fulton/Dekalb counties, 2020.

**Table 4.**
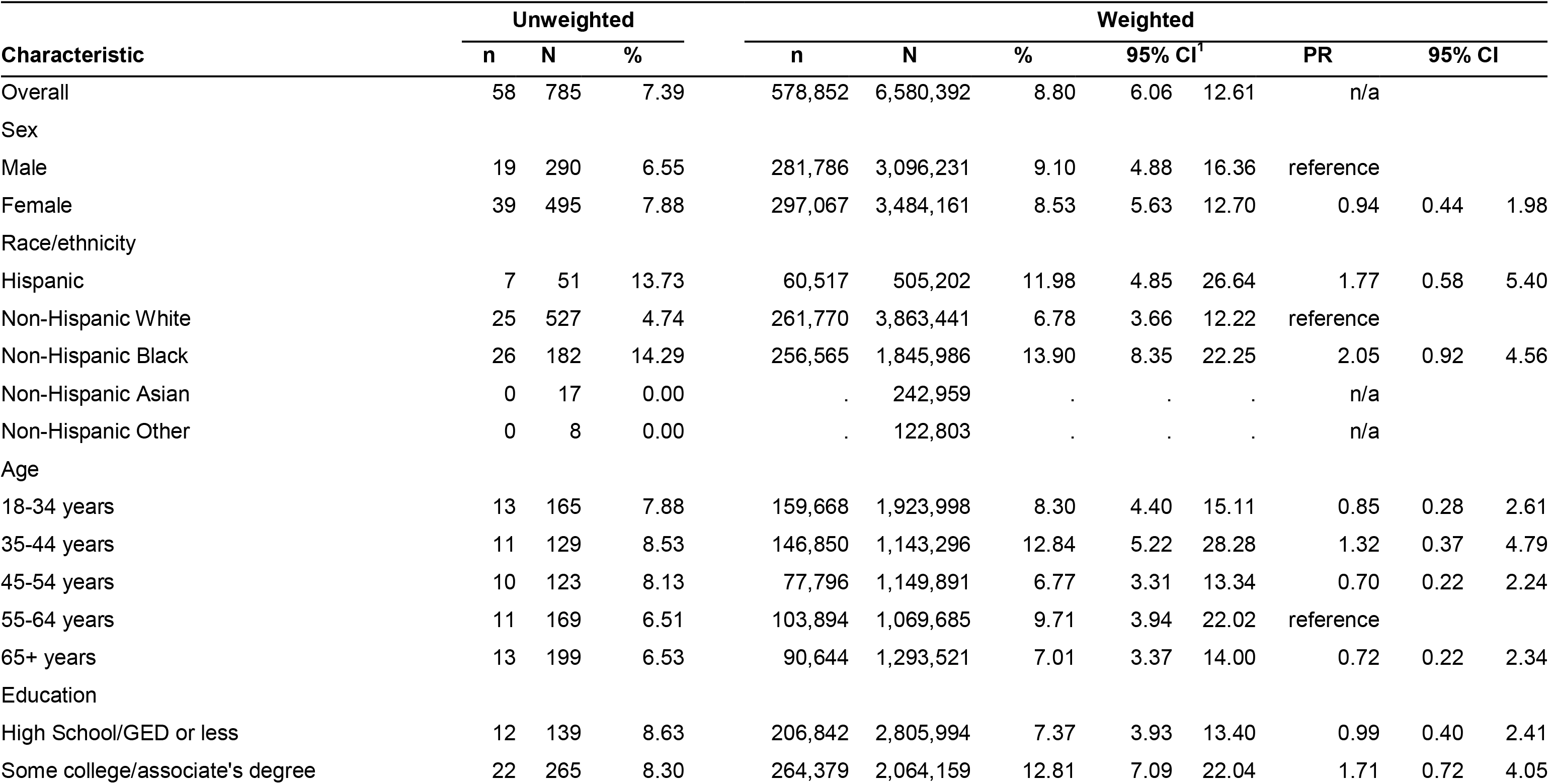
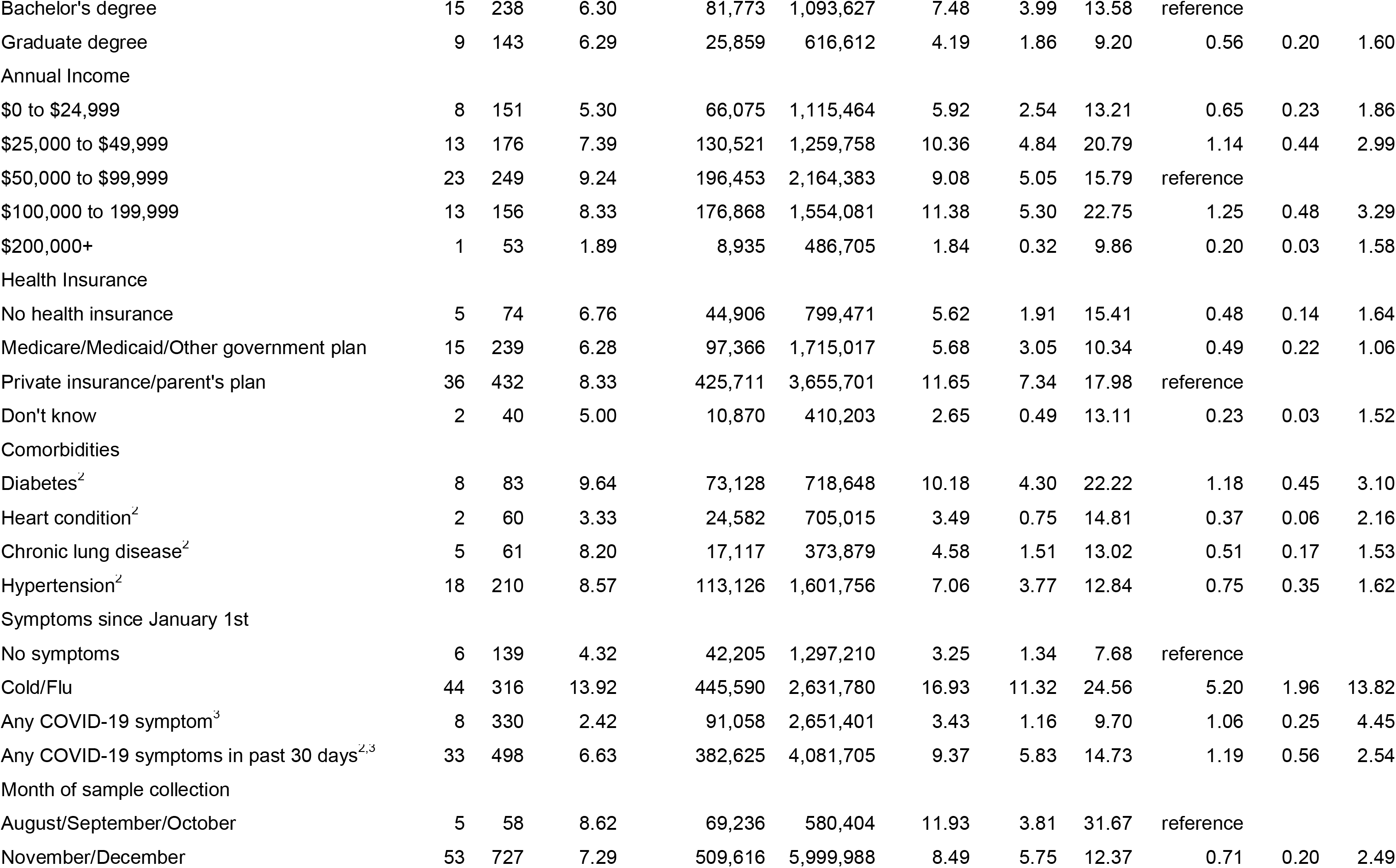
Unweighted and weighted SARS-CoV-2 antibody prevalence for a probability sample of 785 households and weighted results and prevalence ratios, rest of GA outside of Fulton/Dekalb counties, 2020.

| Characteristic | Unweighted |  |  | Weighted |  |  |  |  |  |  |  |
| --- | --- | --- | --- | --- | --- | --- | --- | --- | --- | --- | --- |
|  | n | N | % | n | N | % | 95% CI<sup>†</sup> |  | PR | 95% CI |  |
| Overall | 58 | 785 | 7.39 | 578,852 | 6,580,392 | 8.80 | 6.06 | 12.61 | n/a |  |  |
| Sex |  |  |  |  |  |  |  |  |  |  |  |
| Male | 19 | 290 | 6.55 | 281,786 | 3,096,231 | 9.10 | 4.88 | 16.36 | reference |  |  |
| Female | 39 | 495 | 7.88 | 297,067 | 3,484,161 | 8.53 | 5.63 | 12.70 | 0.94 | 0.44 | 1.98 |
| Race/ethnicity |  |  |  |  |  |  |  |  |  |  |  |
| Hispanic | 7 | 51 | 13.73 | 60,517 | 505,202 | 11.98 | 4.85 | 26.64 | 1.77 | 0.58 | 5.40 |
| Non-Hispanic White | 25 | 527 | 4.74 | 261,770 | 3,863,441 | 6.78 | 3.66 | 12.22 | reference |  |  |
| Non-Hispanic Black | 26 | 182 | 14.29 | 256,565 | 1,845,986 | 13.90 | 8.35 | 22.25 | 2.05 | 0.92 | 4.56 |
| Non-Hispanic Asian | 0 | 17 | 0.00 | . | 242,959 | . | . | . | n/a |  |  |
| Non-Hispanic Other | 0 | 8 | 0.00 | . | 122,803 | . | . | . | n/a |  |  |
| Age |  |  |  |  |  |  |  |  |  |  |  |
| 18-34 years | 13 | 165 | 7.88 | 159,668 | 1,923,998 | 8.30 | 4.40 | 15.11 | 0.85 | 0.28 | 2.61 |
| 35-44 years | 11 | 129 | 8.53 | 146,850 | 1,143,296 | 12.84 | 5.22 | 28.28 | 1.32 | 0.37 | 4.79 |
| 45-54 years | 10 | 123 | 8.13 | 77,796 | 1,149,891 | 6.77 | 3.31 | 13.34 | 0.70 | 0.22 | 2.24 |
| 55-64 years | 11 | 169 | 6.51 | 103,894 | 1,069,685 | 9.71 | 3.94 | 22.02 | reference |  |  |
| 65+ years | 13 | 199 | 6.53 | 90,644 | 1,293,521 | 7.01 | 3.37 | 14.00 | 0.72 | 0.22 | 2.34 |
| Education |  |  |  |  |  |  |  |  |  |  |  |
| High School/GED or less | 12 | 139 | 8.63 | 206,842 | 2,805,994 | 7.37 | 3.93 | 13.40 | 0.99 | 0.40 | 2.41 |
| Some college/associate's degree | 22 | 265 | 8.30 | 264,379 | 2,064,159 | 12.81 | 7.09 | 22.04 | 1.71 | 0.72 | 4.05 |

|  |  |  |  |  |  |  |  |  |  |  |  |  |
| --- | --- | --- | --- | --- | --- | --- | --- | --- | --- | --- | --- | --- |
| Bachelor's degree | 15 | 238 | 6.30 | 81,773 | 1,093,627 | 7.48 | 3.99 | 13.58 | reference |  |  |  |
| Graduate degree | 9 | 143 | 6.29 | 25,859 | 616,612 | 4.19 | 1.86 | 9.20 | 0.56 | 0.20 | 1.60 |  |
| Annual Income |  |  |  |  |  |  |  |  |  |  |  |  |
| \$0 to \$24,999 | 8 | 151 | 5.30 | 66,075 | 1,115,464 | 5.92 | 2.54 | 13.21 | 0.65 | 0.23 | 1.86 | |
| \$25,000 to \$49,999 | 13 | 176 | 7.39 | 130,521 | 1,259,758 | 10.36 | 4.84 | 20.79 | 1.14 | 0.44 | 2.99 | |
| \$50,000 to \$99,999 | 23 | 249 | 9.24 | 196,453 | 2,164,383 | 9.08 | 5.05 | 15.79 | reference | | | |
| \$100,000 to 199,999 | 13 | 156 | 8.33 | 176,868 | 1,554,081 | 11.38 | 5.30 | 22.75 | 1.25 | 0.48 | 3.29 | |
| \$200,000+ | 1 | 53 | 1.89 | 8,935 | 486,705 | 1.84 | 0.32 | 9.86 | 0.20 | 0.03 | 1.58 | |
| Health Insurance |  |  |  |  |  |  |  |  |  |  |  |  |
| No health insurance | 5 | 74 | 6.76 | 44,906 | 799,471 | 5.62 | 1.91 | 15.41 | 0.48 | 0.14 | 1.64 |  |
| Medicare/Medicaid/Other government plan | 15 | 239 | 6.28 | 97,366 | 1,715,017 | 5.68 | 3.05 | 10.34 | 0.49 | 0.22 | 1.06 |  |
| Private insurance/parent's plan | 36 | 432 | 8.33 | 425,711 | 3,655,701 | 11.65 | 7.34 | 17.98 | reference |  |  |  |
| Don't know | 2 | 40 | 5.00 | 10,870 | 410,203 | 2.65 | 0.49 | 13.11 | 0.23 | 0.03 | 1.52 |  |
| Comorbidities |  |  |  |  |  |  |  |  |  |  |  |  |
| Diabetes<sup>2</sup> | 8 | 83 | 9.64 | 73,128 | 718,648 | 10.18 | 4.30 | 22.22 | 1.18 | 0.45 | 3.10 |  |
| Heart condition<sup>2</sup> | 2 | 60 | 3.33 | 24,582 | 705,015 | 3.49 | 0.75 | 14.81 | 0.37 | 0.06 | 2.16 |  |
| Chronic lung disease<sup>2</sup> | 5 | 61 | 8.20 | 17,117 | 373,879 | 4.58 | 1.51 | 13.02 | 0.51 | 0.17 | 1.53 |  |
| Hypertension<sup>2</sup> | 18 | 210 | 8.57 | 113,126 | 1,601,756 | 7.06 | 3.77 | 12.84 | 0.75 | 0.35 | 1.62 |  |
| Symptoms since January 1st |  |  |  |  |  |  |  |  |  |  |  |  |
| No symptoms | 6 | 139 | 4.32 | 42,205 | 1,297,210 | 3.25 | 1.34 | 7.68 | reference |  |  |  |
| Cold/Flu | 44 | 316 | 13.92 | 445,590 | 2,631,780 | 16.93 | 11.32 | 24.56 | 5.20 | 1.96 | 13.82 |  |
| Any COVID-19 symptom<sup>3</sup> | 8 | 330 | 2.42 | 91,058 | 2,651,401 | 3.43 | 1.16 | 9.70 | 1.06 | 0.25 | 4.45 |  |
| Any COVID-19 symptoms in past 30 days<sup>2,3</sup> | 33 | 498 | 6.63 | 382,625 | 4,081,705 | 9.37 | 5.83 | 14.73 | 1.19 | 0.56 | 2.54 |  |
| Month of sample collection |  |  |  |  |  |  |  |  |  |  |  |  |
| August/September/October | 5 | 58 | 8.62 | 69,236 | 580,404 | 11.93 | 3.81 | 31.67 | reference |  |  |  |
| November/December | 53 | 727 | 7.29 | 509,616 | 5,999,988 | 8.49 | 5.75 | 12.37 | 0.71 | 0.20 | 2.49 |  |

### SARS-CoV-2 cumulative incidence

Adjusting estimates for waning detectable antibody levels, the estimated number of cumulative new SARS-CoV-2 infections among Georgian adults was 1,307,518 (95% CrI: 1,081,788-1,541,200) as of November 16, 2020. The cumulative incidence was 16.1% (95% CrI: 13.5-19.2%) (Figure 2). The estimated IFR was 0.78% (95% CrI: 0.66-0.94%). The Georgia Department of Health reported 348,204 COVID-19 cases as of November 16, 2020, indicating that about one-quarter (26.6%; 95% Crl: 22.6%-32.2%) of SARS-CoV-2 infections among adults was reported.

**Figure 2.**
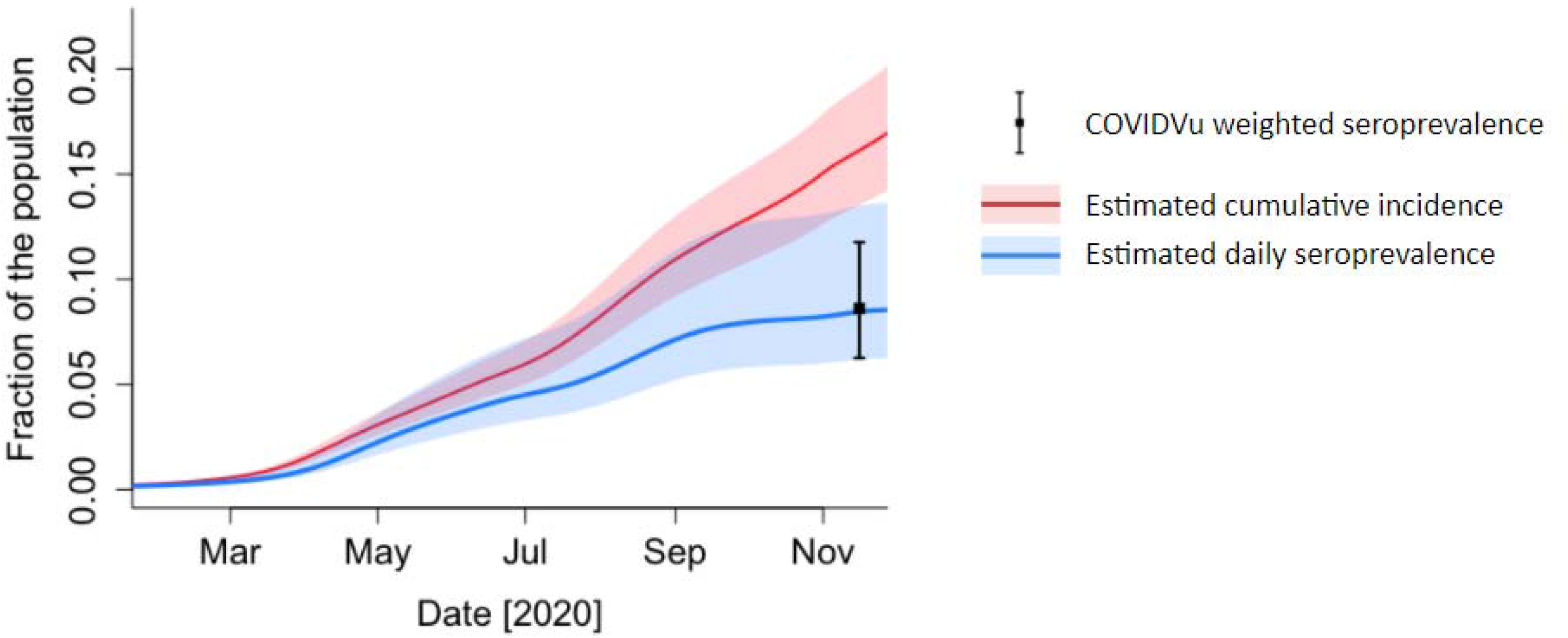
Estimated cumulative incidence of SARS-CoV-2 infection among adults adjusted for waning antibodies and daily seroprevalence, Georgia, 2020.

Among specimens tested with PCR, a total of 16/1,529 (1.0%) were positive. Of these 16, 8 (50.0%) were also reactive for total Ig.

## Discussion

A statewide probability sample of Georgia households conducted between August and December 2020 allowed for robust estimation of the cumulative incidence of SARS-CoV-2 infection among adults, finding that over 16% of Georgia’s adult population - about one in six - had been infected with the virus as of November 2020. Seroprevalence was highest among Hispanic and non-Hispanic Black persons, and similar for the Atlanta-metro counties of Fulton and DeKalb compared to the rest of the state.

The data obtained through this household-based, representative survey complement data on reported COVID-19 cases and overcome key limitations associated with data available through traditional state-based COVID-19 surveillance activities and seroprevalence surveys. Because our household sampling strategy was not restricted to individuals experiencing COVID-19 symptoms or seeking SARS-CoV-2 testing, biases associated with testing availability, test-seeking behaviors and the inability to identify asymptomatic individuals were minimal. Additionally, because these data are obtained from a random, representative sample of Georgia residents, the findings can provide reliable inference to all adult Georgia residents. Due to the finding that, as of mid-November 2020, Georgia had only recognized approximately 26% of adults infected with SARS-CoV-2, there is an ongoing need to lower barriers for testing. Our data also validate efforts made thus far in the pandemic response to encourage and invest in frequent, ample testing, despite pushback by lawmakers and certain segments of the general public who may have viewed public health mitigation strategies to have been excessive.(*16*)

Population-based COVID-19 data at the state level allows for a more nuanced understanding of the continuum of infection, diagnosis and mortality and the relationship of these metrics to programmatic priorities. For example, as of November 16, 2020, Georgia’s estimated case fatality ratio (CFR) was 2.1%.(*17*) Because the CFR is calculated from diagnosed cases (and practically from reported cases), having an IFR (which includes people who were asymptomatic in the denominator) advances our understanding of how common death is among all people infected with COVID-19, regardless of whether those infections were symptomatic. The result is that the CFR overstates how common death is among all those infected in the state. Our more comprehensive IFR that was estimated around the same time was about about a third of the CFR. Accordingly, SARS-CoV-2 infections may not be as fatal as had been previously reported; nonetheless, based on the IFR estimated in this study, 1 out of every 130 adult Georgians who are infected with SARS-CoV-2 will die.

Knowing that Georgia went into its winter surge with 16% of the adult population having had a SARS-CoV-2 infection is informative. It provides a reliable “lower bound” on how many adults have been infected, and despite waning antibodies, the proportion infected will only increase. When coupled with increasing data on duration of immunity, these more robust estimates of cumulative SARS-CoV-2 infection can help decision-makers understand how natural immunity contributes to a Georgia-specific herd immunity metric. To that end, data from our study suggest persons over the age of 65 have experienced far less infection than other age groups (and are therefore still susceptible), validating the state’s decision to prioritize that demographic for vaccination first. Our study also found the highest seroprevalence among Hispanic and non-Hispanic Black persons, suggesting that similar findings from diagnosed cases are not the result of biases in testing. These findings should be used to strengthen messaging around why vaccination remains important for these demographic groups despite previous infection, and add urgency to investments in increasing education and reducing barriers to access for Hispanic and Black Georgians.

Although we lack power to examine differences in seroprevalence by other meaningful geographic units (e.g. health district or state region), the results stratified by Fulton/DeKalb versus the rest of the state offer some useful local insights into differences in infection by metropolitan vs. non-metropolitan areas. Fulton and DeKalb comprise all of the City of Atlanta, Georgia’s capital and most populated city. The populations of these two counties comprise 17% of Georgia’s population.(*18*) Observing that the seroprevalence was similar among residents of those two counties compared to the rest of the state was notable, given that national data show consistently lower diagnosis rates for micropolitan and non-core areas through July 2020, with a switch in the pattern starting in August 2020 such that more infections were reported in less urbanized areas.(*19*) Thus, our finding of similar seroprevalence levels in urban and rural areas of Georgia might represent a combination of more historical infections in urban areas earlier in the year, and a higher concentration of infections in more rural areas during the period of the specimen collection. Infection rates (and subsequently antibody seroprevalence) are also related to risk mitigation behaviors. Although the use of face coverings has always been strongly encouraged across Georgia, the City of Atlanta issued a mask mandate in July 2020. With increasing ecological evidence suggesting the benefits masking can have on reducing community spread of SARS-CoV-2,(*20, 21*) Atlanta’s mask mandate may have limited the propagation of the virus in Fulton and Dekalb counties, where higher levels of transmission might have been favored by higher population density.

Our study is subject to a number of limitations. While we utilized a representative sampling frame, our response rate was 11.3%, which is low but typical for mailed surveys using address-based sampling frames.(*22*) The only other two household samples reported were conducted through door to door offer of enrollment (versus mailout enrollment packages in our study) and also had relatively low response rates (23.6 to 23.7%(*23*)). Our results are likely subject to some degree of differential response bias; we addressed this by oversampling specific groups (e.g., Black and Hispanic households) with lower response rates, and by weighting for non-response of households, a procedure with validity facilitated by the nature of an address-based sampling frame. Importantly, we were only able to address differential non-response using characteristics of the population that were available to us on the frame (e.g., population distributions by race/ethnicity or household income levels). Characteristics that may be associated with COVID-19 risk but not available at the population-level, such as higher general propensity to take risks, were not available for extrapolation to the underlying population from sample data and therefore may contribute to unaddressed selection bias in estimates. Misclassification of antibody status was possible due to waning antibodies(*2, 24*), but unlike other studies reported to date, we accounted for these biases through our modeling approach.(*14*) Our model used an estimated average time of seropositivity from a previous study conducted for New York City (*14*). This estimate was generated for an ELISA kit against the SARS-CoV-2 spike protein that detects the total immunoglobulin response (*25*), which is different from what we used in this study, but this is the only estimate of the timeline of the population-level waning antibody available at this point.

Knowing the true proportion of people who have been previously infected with SARS-CoV-2 is useful both epidemiologically and practically. Past seroprevalence studies from convenience samples and biased samples of residual blood provide important information, but the results are subject to selection biases associated with the sources of specimens. For Georgia, having reliable estimates of the cumulative incidence of SARS-CoV-2 infection among adults allows for more informed decision-making about risk mitigation and vaccination strategies. Data collections will be repeated in March and June of 2021, and results will be examined in an ongoing way as knowledge advances on topics ranging from duration of immunity to implications of antibodies for protection against novel variants.

## Data Availability

COVIDVu study data are not yet publicly available. Data used for the ratio of the cases reported to cumulative incidence cases (the reported fraction) was developed from confirmed PCR+ cases in Georgia as of November 16, 2020 using data for adults ≥18 years from the Georgia Department of Public Health's public use dataset: https://ga-covid19.ondemand.sas.com/docs/ga_covid_data.zip

## Acknowledgements

The authors thank Salesforce for its donation of Salesforce.org licenses and system development for the project, and to acknowledge the contributions of development team members Lee Evans, John Thrasher, Stephen Noe, Anurag Jaiswal, Ryan Williams, David Affentranger, and Todd Siegler. They also thank the Kaiser Family Foundation for their design and thoughtful contributions.

